# Analysis of SARS-CoV-2 mutations associated with resistance to therapeutic monoclonal antibodies that emerge after treatment

**DOI:** 10.1101/2023.03.02.23286677

**Authors:** Daniele Focosi, Scott McConnell, David J. Sullivan, Arturo Casadevall

**Affiliations:** North-Western Tuscany Blood Bank, Pisa University Hospital, Italy; Department of Molecular Microbiology and Immunology, Johns Hopkins Bloomberg School of Public Health, Baltimore, MD 21205, USA

**Keywords:** COVID-19, SARS-CoV-2, monoclonal antibodies, sotrovimab, casirivimab, imdevimab, tixagevimab, cilgavimab

## Abstract

The mutation rate of the Omicron sublineage has led to baseline resistance against all previously authorized anti-Spike monoclonal antibodies (mAbs). Nevertheless, in case more antiviral mAbs will be authorized in the future, it is relevant to understand how frequently treatment-emergent resistance has emerged so far, under different combinations and in different patient subgroups. We report the results of a systematic review of the medical literature for case reports and case series for treatment-emergent immune escape, which is defined as emergence of a resistance-driving mutation in at least 20% of sequences in a given host at a given timepoint. We identified 31 publications detailing 201 cases that included different variants of concern (VOC) and found that the incidence of treatment emergent-resistance ranged from 10% to 50%. Most of the treatment-emergent resistance events occurred in immunocompromised patients. Interestingly, resistance also emerged against cocktails of two mAbs, albeit at lower frequencies. The heterogenous therapeutic management of those cases doesn’t allow inferences about the clinical outcome in patients with treatment-emergent resistance. Furthermore, we noted a temporal correlation between the introduction of mAb therapies and a subsequent increase in SARS-CoV-2 sequences across the globe carrying mutations conferring resistance to that mAb, raising concern as to whether these had originated in mAb-treated individuals. Our findings confirm that treatment-emergent immune escape to anti-Spike mAbs represents a frequent and concerning phenomenon and suggests that these are associated with mAb use in immunosuppressed hosts.

## Introduction

Passive antibody pre-exposure and post-exposure prophylaxis and treatments are much needed for immunocompromised patients at risk of COVID-19 progression. For them, anti-Spike monoclonal antibodies (mAb) have been the treatment of choice since early 2021. The biologicals were safe, well tolerated and efficient at preventing disease and reducing the likelihood of progression to severe disease. Unfortunately, the advent of the Omicron VOC progressively defeated the emergency use authorized mAbs, leaving all revoked by the US Food and Drug Administration at the time of writing. The COVID-19 pandemic has represented the first instance of large-scale deployments of passive immunotherapies against an infectious agent, and with persistence at epidemic levels and fluctuations in the dominant variants, it is possible that this scenario will persist for more years. This in turn provides a unique opportunity to study the phenomenon of emerging resistance to antibody-based therapeutics. The emergence of mAb-resistant variants after individualized therapy has potential public health implications if these viruses re-enter the general population and are able to evade vaccine or prior infection immunity by virtue of their mutations (Casadevall and Focosi, 2023)Here we analyzed mAbs resistance patterns following therapy principally in immunosuppressed but sometimes in immunocompetent COVID-19 patients.

## Materials and methods

### Inferring baseline resistance to anti-Spike monoclonal antibodies

The Stanford University Coronavirus Antiviral & Resistance Database is a comprehensively curated collection of published data on the susceptibility of SARS-CoV-2 variants to anti-Spike mAbs and the plasma from previously infected and/or vaccinated persons. It also records the Spike protein mutations that are selected by mAbs and that emerge in persons experiencing prolonged infection (Tzou et al., 2020). It is publicly accessible at https://covdb.stanford.edu/. For each authorized anti-Spike mAb, we manually scanned mutations associated with >5-fold median reduction in neutralizing antibody (nAb) titers from primary research listed in search results provided at https://covdb.stanford.edu/search-drdb/?form_only. The cutoff was arbitrarily chosen by the authors based on former literature suggesting loss of therapeutic efficacy.

### Literature search

On February 1 2023, we searched PubMed, bioRxiv and medRxiv repositories for literature published since January 1 2020 to February 1 2023 using English language as a restriction. We used the query “(“sotrovimab” OR “tixagevimab” OR “cilgavimab” OR “bamlanivimab” OR “etesevimab” OR “casirivimab” OR “imdevimab” OR “bebtelovimab” OR “regdanvimab”) AND (“resistance” OR “escape” OR “evasion” OR “evolution”)”. The PRISMA flowchart is reported in Figure 1. We excluded all primary research on baseline (as opposed to treatment-emergent) resistance and all secondary research. We collected data about sample size, underlying immunosuppression and incidence of *de novo* mutations at Spike residues associated with mAb escape.

**Figure 1.**
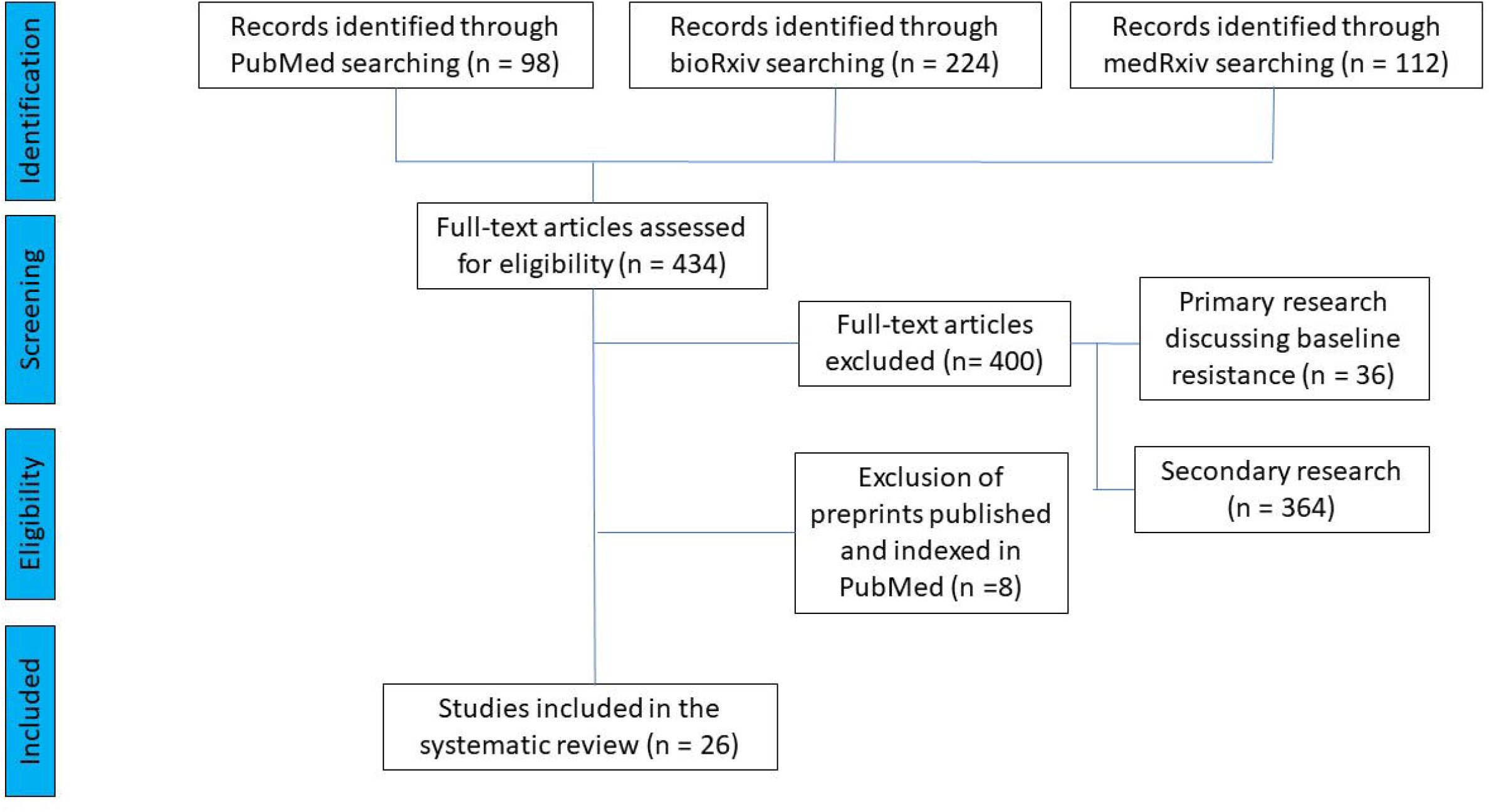
PRISMA flowchart for study selection.

### Determining the temporal relationship between mAb authorization and global prevalence

On February 1 2023 we searched CoV-Spectrum.org (Chen et al., 2021a) for worldwide prevalence of SARS-CoV-2 sublineages carrying either at least one mutation conferring resistance to a given mAb or just the major resistance associated mutations (i.e., Q493X for bamlanivimab and etesevimab, R346X for cilgavimab, K444X for bebtelovimab).

### Mapping treatment emergent mutations to the structures of Spike:mAb complexes

All three-dimensional molecular representations were generated with PyMOL 2.5.2 (Schrodinger). Deposited structures of therapeutic mAbs in complex with the Spike protein were obtained from the Protein Database (PDB) under the accession numbers 7KMG (bamlanivimab), 7C01 (etesevimab), 6XDG (casirivimab/imdevimab), 7L7E (cilgavimab), 7L7D (tixagevimab) and 6WPS (sotrovimab).

## Results

The individual Spike amino acid mutations conferring resistance to each mAb are summarized in Table 1. A review of 31 publications reporting mAb resistance yielded a list of Spike protein mutations associated with mAb resistance (Table 2). From these publications we identified 201 out of 1060 patients who experienced emergence of resistant SARS-CoV-2 after treatment with one of the anti-Spike mAb. As expected from real-world indications and usage, the majority of these patients were immunocompromised, but immunocompetent patients were also noted (Sabin et al., 2022).

**Table 1.**
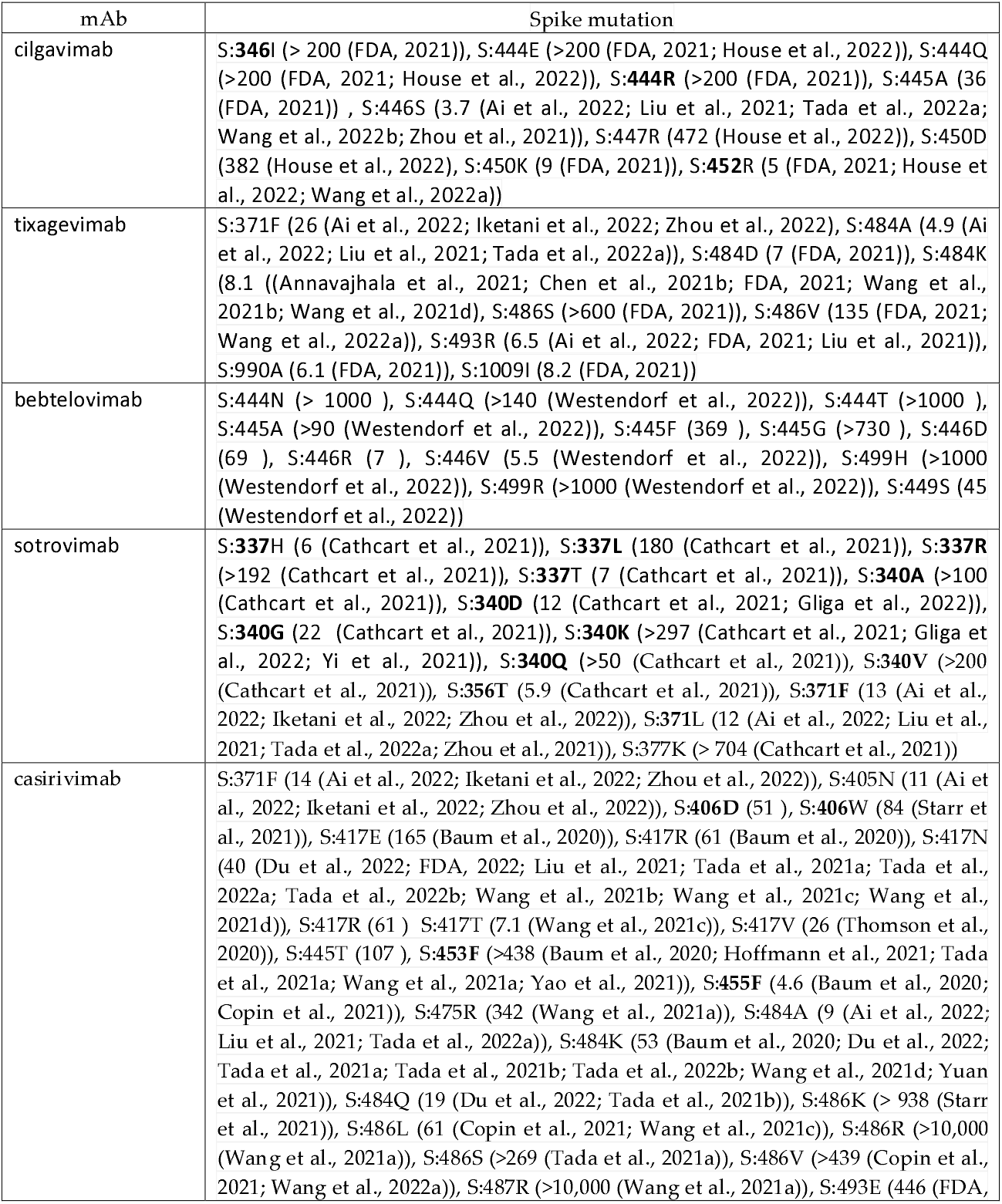
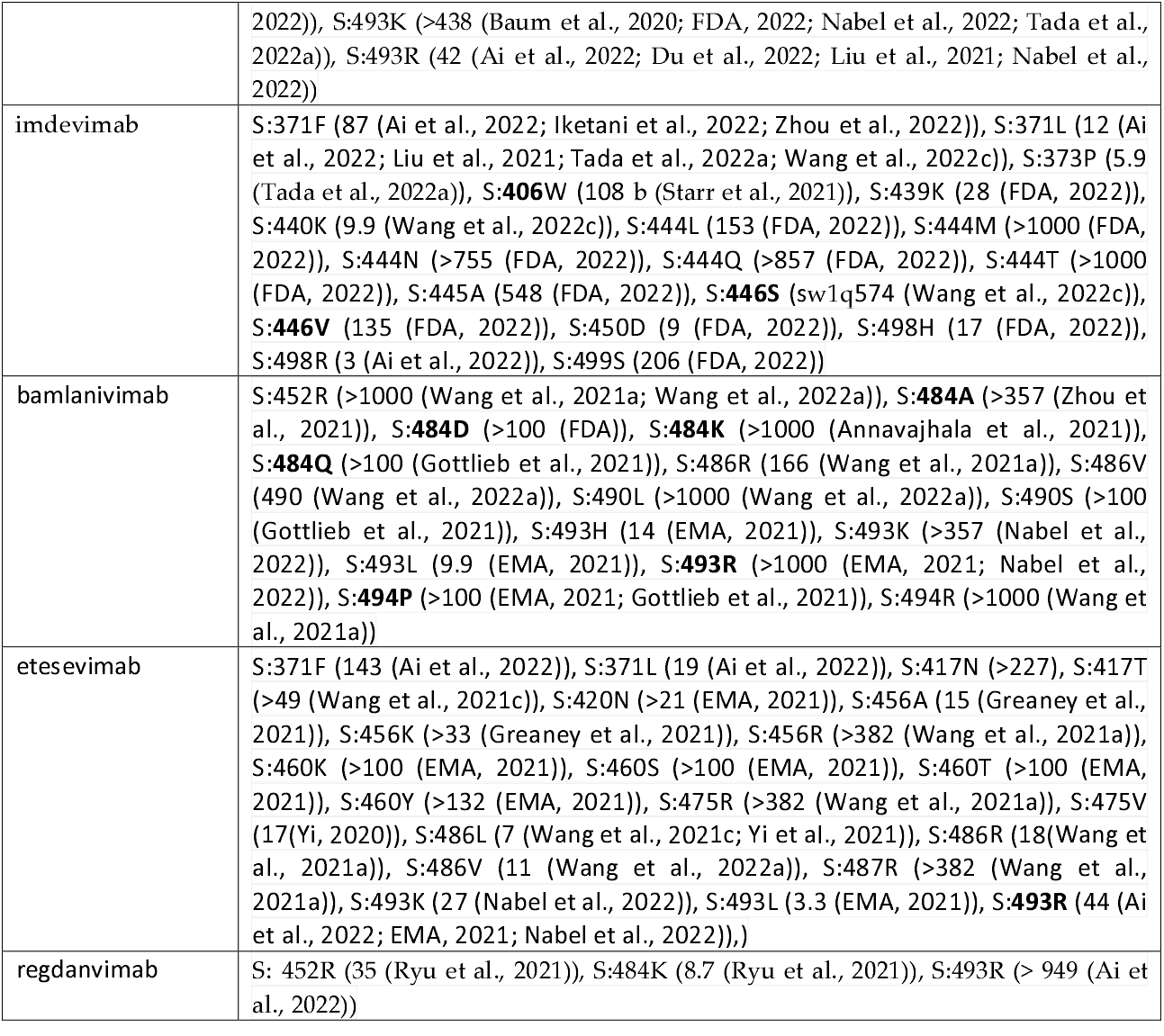
Spike mutations associated with mAb resistance in *in vitro* studies. Bold characters show the ones that have been detected as emerged *in vivo* after treatment with the specific mAb. In cases where the exact amino acid change has not been studied *in vitro*, only the residue is highlighted. Number within parentheses represent the median fold-reduction in neutralizing antibody titers.

**Table 2.** Published reports of anti-Spike mAb treatment-emergent resistance.

| mAb(s) | N (of n) cases | SARS-CoV-2 strain | Spike mutations | Ref |
| --- | --- | --- | --- | --- |
| bamlanivimab monotherapy (700 mg iv) | 7 out of 101 (6.9%) immunocompetent patients | n.a. | E484K/Q, S494P | Choudhary *et al* (Boucau et al., 2022; Choudhary et al., 2022) |
|  | 1 immunocompromised patient | Alpha | E484K, Q493R | Truffot *et al* (Truffot et al., 2021) |
|  | 1 immunocompromised patient | Alpha | E484K, Q493R, S494P | Lohr *et al* (Lohr et al., 2021) |
|  | 5 out of 6 (83.3%) immunocompromised patients | B.1 | E484K → E484Q, reverted to E484K after CCP | Jensen *et al* (Jensen et al., 2021) |
|  | 1 immunocompromised patient | Alpha | E484Q | Bronstein *et al* (Bronstein et al., 2021) |
|  | 2 immunocompromised patients | 20B/20G | E484Q/K | Simons *et al* (Simons et al., 2022) |
|  | 1 immunocompetent patient | B.1.311 | E484K | Sabin *et al* (Sabin et al., 2022) |
|  | 1 immunocompromised patient | B.1.2 | E484T | Halfmann *et al* (Halfmann et al., 2022) |
|  | 2 immunocompromised patients | 20A.EU2 / 20I/501Y.V1 | E484A/K | Destras *et al* (Destras et al., 2021) |
|  | 2 immunocompromised patients |  | E484Q/K, Q493R | Scherer et al (Scherer et al., 2022) |
|  | 5 out of 45 (11.1%) | Alpha | E484A/K, Q493R, S494P | Gupta *et al* (Gupta et al., 2023) |
|  | 1 out of 2 (50%) immunocompetent | B.1.214.2 Alpha | Q493R | Jary *et al* (Jary et al., 2022) |
|  | 5 out of 6 (83.3%) | Alpha | E484A/K, Q493R, S494P | Peiffer-Smadja *et al* (Peiffer-Smadja et al., 2021) |
| bamlanivimab 700 mg + etesevimab 1400 mg cocktail | 1 out of 102 (1%) patients | B.* | S494P | Gottlieb *et al* (Gottlieb et al., 2021) |
|  | 1 immunocompromised patient | Alpha | Q493R | Focosi *et al* (Focosi et al., 2021) |
|  | 1 immunocompromised patient |  | Q493R | Guigon *et al* (Guigon et al., 2021) |
|  | 5 out of 23 (21.7%) patients (17 immunocompromised) |  | E484K, Q493R | Vellas *et al* (Vellas et al., 2021) |
|  | 3 out of 108 (2.8%) |  | E484D, Q493R/K | Gupta *et al* (Gupta et al., 2023) |
|  | 1 out of 3 (33%) |  | Q493R | Jary *et al* (Jary et al., 2022) |
|  | 5 out of 34 (14.7) patients |  | Q493R, E484D | Pommeret *et al* |
|  | (5 with B-cell malignancies) |  |  | (Pommeret et al., 2021) |
| casirivimab +<br>imdevimab<br>(REGN-CoV)<br>cocktail | 1 immunocompromised patient | n.a. | E484K/A, Y489H, Q493K and N501Y | Choi *et al* (Choi et al., 2020)<br>Clarke *et al* (Clark et al., 2021) |
|  | 0 out of 8 (0%) | Alpha (7) and Gamma (1) | - | Jary *et al* (Jary et al., 2022) |
|  | 1 out of 17 (5.9%) | Alpha | E406G | Gupta *et al* (Gupta et al., 2023) |
|  | 15 out of 160 (9.4%) | Delta, BA.1 | E406D/Q, G446S/V, Y453F, L455F/S | Ragonnet-Cronin *et al* (Ragonnet-Cronin et al., 2022) |
| sotrovimab | 4 cases out of 100 (4%) | Delta | E340K/A/V | Rockett *et al* (Rockett et al., 2021) |
|  | 10 cases out of 18 (55.6%)<br>(15 immunocompromised) | BA.1 (94%)<br>BA.2 (6%) | E340K/A/V/D/G/Q, P337L/R/S | Birnie *et al* (Birnie et al., 2022) |
|  | 3 out of 16 (18.8%) immunocompromised patients | 2 BA.1, 1 BA.2 | E340D | Focosi *et al* (Focosi et al., 2022b) |
|  | 18 out of 34 (52.9%) immunocompromised patients | 17 BA.1<br>1 BA.2 | P337L/S, E340A/K/D/G, K356T, S371F | Vellas *et al* (Vellas et al., 2022b) |
|  | 4 of 25 (16%) BA.1-infected patients<br>2 of 7 (28.6%) BA.2-infected | BA.1<br>BA.2 | P337X, E340X | Huygens *et al* (Huygens et al., 2022) |
|  | 5 out of 8 (62.5%) immunocompromised | BA.1 (7)<br>AY.100 (1) | P337L, E340D/R/K/V/Q, R346T, K356T | Andrés *et al* (Andrés et al., 2023) |
|  | 8 patients | BA.1 | P337R/S, E340A/D/K/Q | Destras *et al* (Destras et al., 2022) |
|  | 9 out of 34 (26.5%) | BA.1.1.* (n=14)<br>BA.1 (n = 13)<br>BA.2 (n = 7) | E340K/D/V | Gupta *et al* (Gupta et al., 2023) |
|  | 54 out of 134 (40.3%) patients | Delta, BA.1, BA.2 | P337R/S, E340A/D/K/V, K356T | Ragonnet-Cronin *et al* (Ragonnet-Cronin et al., 2022) |
|  | 14 out of 43 (32.6%) patients | BA.1 | P337S/H/L/R, E340K/D/V | Gliga *et al* (Gliga et al., 2022) |
| cilgavimab-tixagevimab | 9 out of 18 (50%) immunocompromised cases | BA.2 | K444R/N, R346T, L452M | Vellas *et al* (Vellas et al., 2022a) |
|  | 1 out 19 (5.3%) immunocompetent cases | BA.4/5 | K444N | Maggi *et al* (Fabeni et al., 2023) |
| bebtelovimab | 1 immunocompromised patient | BA.1.23 | V445A | Gonzalez-Reiche *et al*<br>(Gonzalez-Reiche et al., 2022) |

26 publications reported incidence rates while investigating a cohort: among them the incidences varied between 0% to 100% with overall 195/1079 or 18.1%. In particular bamlanivimab (n=5) ranged from 11 to 83%, sotrovimab (n=10) from 16 to 100%, while dual mAbs (n=10) ranged from 0% to 50%.

The majority of resistant cases from cohorts were reported after mAb monotherapies bamlanivimab (23/160-14%) or sotrovimab (131/427-31%) as opposed to mAb cocktails bamlanivimab+ etesevimab (15/270-6%) or casirivimab+ imdevimab (16/185-9%) (Table 3). The cilgavimab+ tixagevimab essentially had a single active mAb during most of its deployment and the rate in the series cohorts was similar to single monoclonals (10/37-27%).

**Table 3.** Frequencies of treatment-emergentresistance.

| mAb (n=number of cohort series) | # of patients developing treatment-emergent resistance mutations across all case series | total # of patients across all case series | percent of patients developing treatment-emergent resistance across all case series | number of additional single case reports |
| --- | --- | --- | --- | --- |
| bamlanivimab (n=5) | 23 | 160 | 14.4% | 11 |
| bamlanivimab+ etesevimab (n=5) | 15 | 270 | 5.6% | 2 |
| casirivimab+ imdevimab (n=3) | 16 | 185 | 8.6% | 1 |
| sotrovimab (n=11) | 131 | 427 | 30.7% | 0 |
| cilgavimab-tixagevimab (n=2) | 10 | 37 | 27.0% | 0 |
| bebtelovimab (n=0) | n.a. | n.a. | n.a. | 1 |
| Total | 195 | 1079 | 18.1 | 15 |

Studies with a specific mAb or mAb cocktail largely reported the same set of emerging Spike mutations (e.g., Q493X and E484X for bamlanivimab, P337X and E340X for sotrovimab).

By querying the Spike protein sequence database for the major mutations associated with mAb-resistance we noted that these mutations emerged in the general population across the globe shortly after marketing authorization of the mAbs to whom they confer resistance (Figure 3). The most striking examples are the emergence of S:Q493R (the single mutation conferring resistance to both bamlanivimab and etesevimab, in addition to casirivimab and regdanvimab) in Omicron BA.1 shortly after the massive deployment of bamlanivimab/etesevimab cocktail, and the emergence of S:R346X in many converging Omicron sublineages shortly after the massive deployment of cilgavimab for post-exposure prophylaxis in immunosuppressed patients. There was a single case report of bebtelovimab resistance (S:V445A post treatment in an immunocompromised patient with the B.1.23 lineage.

## Discussion

The emergence of mAb treatment resistance during therapy is a common and consistent phenomenon. The discrepancy between the diversity of mutations reported *in vitro* (Table 1) and the few ones detected *in vivo* (Table 2) can be explained by various factors. First, not all mutants selected *in vitro* are fit enough to compete within the intrahost quasispecies swarm. Fitness would include the ability to bind to its cellular receptor as well capacity for virion stability and assembly. The large number of mutations shown to confer resistance *in vitro* that are not reflected in the clinical data could be rationalized by the fact that a reduction in nAb titers measured *in vitro* reflects a defect in antibody binding to Spike variants but does not report on the relative efficacy of ACE2 binding compared to the ancestral Spike. Second, in many cases some mutations were already occurring at baseline (e.g., F486X in patients treated with tixagevimab). Third, among the residues wild-type at baseline, the mutations associated with the highest fold-reduction in neutralization have been more commonly reported: while this could be a notoriety bias by the investigators (placing more attention to the mutations which have been previously reported), it is likely that as soon as one of the highly resistant mutations emerges, emergence of additional mutations do not provide any further fitness advantage.

No cases of treatment-emergent resistance have been reported in the medical literature after regdanvimab: this is likely due to usage much limited in time and locations across the globe. Regdanvimab is not effective against Omicron lineages, limiting its use toa few weeks in the end 2021.

Most anti-Spike mAbs authorized thus far were not formally investigated in randomized controlled trials (RCT) involving immunocompromised patients prior to deployment (Focosi et al., 2022a). Immunocompromised patients are at increased risk for anti-Spike mAb treatment-emergent resistance for multiple risk factors: higher basal viral load, higher likelihood of viral persistence because of inadequate immune response, and contraindications to other directly-acting antivirals (Casadevall and Focosi, 2023). Resistance monitoring is not routinely performed after outpatient mAb treatment, and viral sequencing interpretation requires paying attention to minoritarian sequences within the quasi-species swarm (Vellas et al., 2022a).

Despite FDA deauthorization, many US physicians continued prescription of deauthorized anti-Spike mAbs(Anderson et al., 2022), mostly based on personal beliefs in discrepancies between *in vitro* and *in vivo* activities. The latter phenomenon has been much amplified across Europe (Focosi and Tuccori, 2022), where EMA has never withdrew a single authorized mAb to date, and only sporadically issued alerts about possible basal resistance. In addition, a plethora of uncontrolled or poorly controlled literature is advocating continued efficacy of anti-Spike mAbs based on marginal gains in surrogate endpoints in non-RCTs (Harman et al., 2022) or no change in hospitalization rates of outpatients (Patel et al., 2023). While binding affinity seems a robust surrogate endpoint, advocates of residual *in vivo* activity argue that the concentrations achieved by mAbs *in vivo* can overcome several degrees of baseline resistance to neutralization or that alternative effector function still persists(Stadler et al., 2022a; Stadler et al., 2022b; Wu et al., 2022). Both arguments do not seem robust: with IC_50_ above 1,000 (Cao et al., 2022) antigen binding by the mAb is compromised, and effector functions other than neutralization invariably require binding to the virus to activate Fc-receptors. Under the current variant soup (Focosi et al., 2023), the incompatible turnaround time of viral sequencing and therapeutics delivery, tools to predict basal efficacy of anti-Spike mAbs based on regional genomic surveillance have been developed (Focosi, 2023).

Of interest, several cases of mAb-treatment-emergent immune escape were rescued with COVID-19 convalescent plasma (Halfmann et al., 2022; Pommeret et al., 2021), which, in contrast to mAbs, is polyclonal and thus more difficult to defeat by viral evolution.

We note that in many patients the emergence of mAb resistance was associated with only one amino acid substitution in the epitope, establishing the high vulnerability of monotherapy for selecting resistant variants. Emergence of resistance requires selection of mutant viruses that are fit for replication and that could explain the repeated independent isolations of variants with mutations at specific residues, such as in position 484 with bamlanivimab monotherapy. For some mAbs, the generated footprint is so unique (e.g. S:340X after sotrovimab) that baseline sequencing is not needed to confirm treatment emergence (S:340X being exceedingly rare in GISAID).The transmissibility of such mAb treatment-generated lineages remains a major concern, and it has been formally demonstrated in at least one study (Sabin et al., 2022). Clinical reports of treatment-emergent resistance in response to therapeutic mAbs reviewed here encompass nearly every epitope of the receptor binding domain (RBD), including all 3 receptor binding motif epitopes and the cross reactive S309 site (Figure 2F). Despite partial or complete overlap of antibody epitope on RBD with the ACE2 interface, these positions demonstrate remarkable mutability while maintaining receptor affinity.

**Figure 2.**
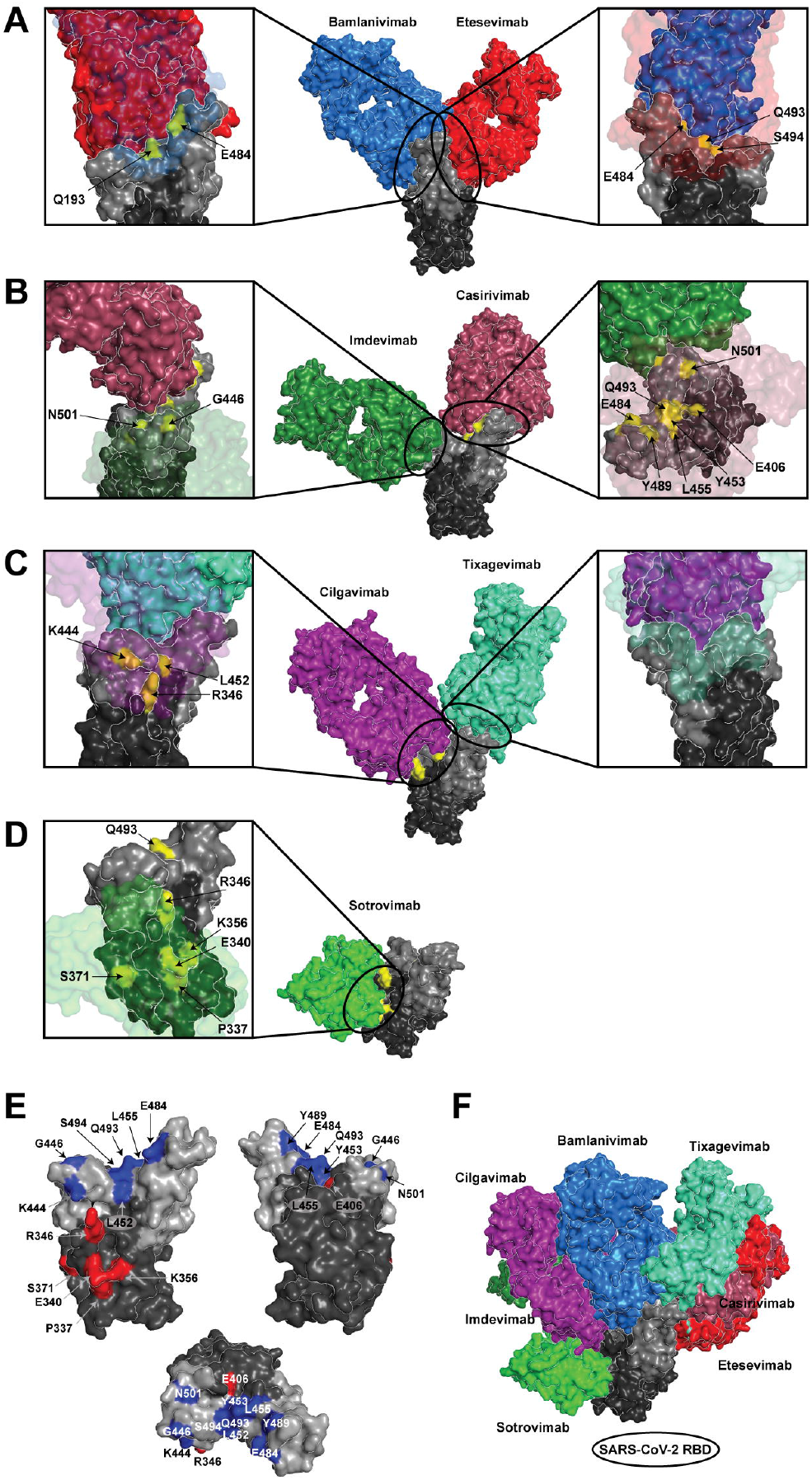
Resistance mutations on the Spike receptor binding domain (RBD) reported after treatment with authorized anti-Spike mAbs. A-D) The complex structures of RBD and corresponding therapeutic mAbs are represented as space-filling surfaces, where the RBD is colored black, the receptor binding motif (residues 438-507) is colored dark gray, and corresponding escape mutations are highlighted in yellow. In the middle panel, the entire RBD-mAb complex is depicted, and expanded and rotated views of each interface are presented in boxes on each side of the main complex. The interacting mAb is partially transparent to enable visualization of the full RBD paratope and reported resistance mutations are labeled. A) The structural complex of the bamlanivimab/etesevimab mAb cocktail with RBD is depicted. Bamlanivimab and etesevimab (PDBID 7KMG, 7C01) are colored blue and red, respectively. B) The structural complex of the casirivimab/imdevimab mAb cocktail (REGN-CoV) with RBD is depicted. Casirivimab and imdevimab (PDBID 6XDG) are colored salmon and dark green, respectively. C) The structural complex of the cilgavimab/tixagevimab mAb cocktail with RBD is depicted. Cilgavimab and tixagevimab (PDBID 7L7E, 7L7D) are colored purple and cyan, respectively. D) The structural complex of the sotrovimab mAb monotherapy with RBD is depicted. Sotrovimab (PDBID 6WPS) is colored light green. E) A summary of the reported treatment-emergent resistance mutations on RBD across all approved anti-Spike therapeutic mAbs are highlighted on the structure of RBD, shown with the front face, back face and top RBM view. Escape mutations in the RBM are highlighted in blue, non-RBM mutations are colored red. F) The complexes of the each of the mAbs in complex with RBD are depicted, colored as in panels A-D, to illustrate the relative binding angles and epitopes of each mAb.

**Figure 3.**
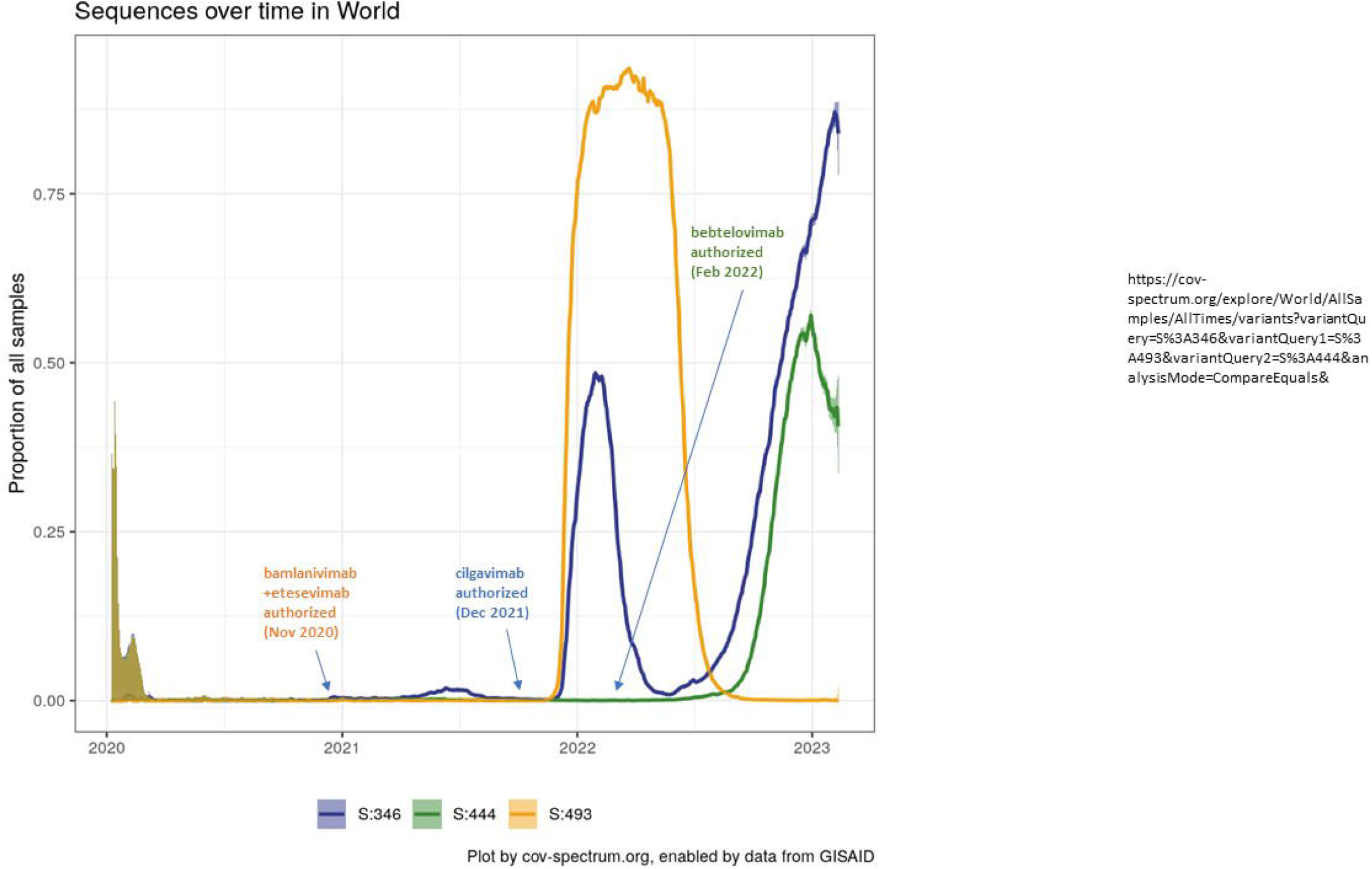
Timeline of anti-Spike mAb authorizations and emergence of key Spike mutations associated with mAb resistance. Figure generated using Cov-Spectrum.org (Chen et al., 2021a)

Since therapeutic mAbs ultimately trace their ancestry to immune B cells in humans, the emergence of mAb resistance mutations poses the theoretical concern that once these occur in immunosuppressed individuals the mAb-resistant variants can spread to immunocompetent populations, bypass existing immunity, and create new waves of infection (Casadevall and Focosi, 2023). Consistent with this concern we found a temporal association between the introduction of the mAb and an increase in frequency in the SARS-CoV-2 Spike protein sequence database. Such temporal association could be explained by a scenario whereby the emergence of these variants in immunosuppressed hosts was followed by their spread among the general population. Such mutations would have created antigenically distinct viruses that could have an advantage in bypassing established immunity in the general population from vaccination and prior infection as these occur in a critical neutralizing epitope. We caution that association does not imply causation and that other interpretations are possible. For example, immunity to previous infection waves could be another possible driver for selection of novel Spike variants, and the possibility of viral transitioning through animal reservoirs remains as possibility since Q493R is a mouse-adapting mutation, which in turn suggests the possibility of reverse zoonosis. Nevertheless, the temporal association between mAb introduction in clinical practice and increased frequency of mutations conferring resistance to that mAb is concerning and suggest that future use of mAb therapies in immunocompromised populations should consider the use of cocktails and/or combination therapy with small molecule antivirals. Cocktails-of-three or more anti-Spike mAbs with non-overlapping epitopes could minimize the chances for immune escape.

## Data Availability

All the data used for this manuscript are available on PubMed,bioRxiv and medrXiv as per Figure 1

